# Modeling the Effects of Formulary Exclusions: How Many Patients Could Be Affected by a Specific Exclusion?

**DOI:** 10.1101/2023.10.31.23297782

**Authors:** Anne M. Sydor, Emily Bergin, Jonathan Kay, Erik Stone, Robert Popovian

## Abstract

**Background:** Drug formularies, initially designed to promote the use of cost-effective generic drugs are now designed to maximize financial benefits for the pharmacy benefit management companies that negotiate drug purchase prices. In the largest publicly available formulary, 55% of mandated substitutions are not for generic versions of the same active ingredient and/or formulation and may not be medically or financially beneficial to patients.

**Methods:** We modeled the effect of excluding novel agents for atrial fibrillation/venous thromboembolism, migraine prevention, and psoriasis, which all would require substitution with a different active ingredient. Using population data, market share of the two largest U.S. formularies, and 2021 prescription data, we calculated how many people could be affected by such exclusions. Using data from the published literature, we calculated how many of those individuals are likely to discontinue treatment and/or have adverse events due to a formulary exclusion.

**Results:** The number of people likely to have adverse events due to the exclusion could be as high as one million for atrial fibrillation/venous thromboembolism, 900,000 for migraine prevention, and 500,000 for psoriasis. The numbers likely to discontinue treatment for their condition are as high as 924,000 for atrial fibrillation/venous thromboembolism, 646,000 for migraine, and 138,000 for psoriasis.

**Conclusion:** Substitution with a nonequivalent treatment is common in formularies currently in use and is not without substantial consequences for hundreds of thousands of patients. Forced drug substitution results in costly increases in morbidity and mortality and should be part of the cost-benefit analysis of any formulary exclusion.

## BACKGROUND

### Formularies

Formularies developed by insurers and pharmacy benefit management companies (PBM) were originally intended to promote the use of cost-effective treatment, prioritizing less costly medications.^1^ In effect, formularies increased the utilization of generic equivalent drugs.^2^ A generic equivalent is a medication with the same active ingredient and formulation as the patented brand-name medication, intended to have the same clinical effect for the patient as the brand-name drug.^3^

Today, formularies are constructed to limit the number of medicines available for patients based on financial gains for the PBMs.^4^ Hence, many mandated formulary exclusions or substitutions are not necessarily equivalent.^5^ Also, it is impossible to determine whether formulary changes are less costly for patients, employers, or the government because the list price of medication is not the price that a PBM or an insurer pays.^6^ Instead, insurers and PBMs negotiate steep, non-transparent concessions, lowering the net price for only the PBMs and insurers. Unfortunately for patients, the list price determines their out-of-pocket costs related to co-insurance and deductibles.^7^ Less than 1% of the concessions received by PBMs and insurers are passed on to the patients.^8^

From a therapeutic standpoint, the substitutions and exclusions often are not medically beneficial or appropriate for most patients. Our previous research, published in 2022, demonstrated that almost half of all exclusions mandated by the second largest PBM in the U.S. marketplace, ESI, were non-equivalent. Such exclusions force patients to use medications with a different active ingredient, formulation, or mode of administration than their clinician had prescribed.^5^ In some cases, medications were excluded with no alternative, effectively denying patients any treatment. In our updated 2023 research, the number of exclusions by ESI with questionable medical and economic benefits to patients rose to more than half (57.4%).^9^

This current study assesses PBM formulary exclusions by drug class and indication. We intend to estimate the number of patients affected if a specific cardiovascular, dermatologic, or neurologic drug was excluded from ESI or CVS Health formularies in the U.S.

### The Number of People in the U.**S. with Commercial and Medicare Part-D Pharmacy Benefits Provided through PBMs**

In 2022, the U.S. Census Bureau estimated the U.S. population as 333.3 million individuals.^10^ Data from the Kaiser Family Foundation (KFF) for 2021 showed that 48.5% of the U.S. population (∼161.6 million people) had private insurance through their employer, 6.1% (∼20.3 million people) had small group private insurance, and 8.6% (∼28.7 million people) had no health insurance.^10,11^ Another KFF report showed that 14.7% of the U.S. population (∼48.9 million) had Medicare Part D coverage.^12^ Taken together, these numbers suggest that 69.3% of the U.S. population (∼230.9 million people) were covered by commercial or Medicare Part D formularies, with the remaining 22.1% of the U.S. population (∼73.7 million) covered by government insurance (Figure 1).

**Figure 1.**
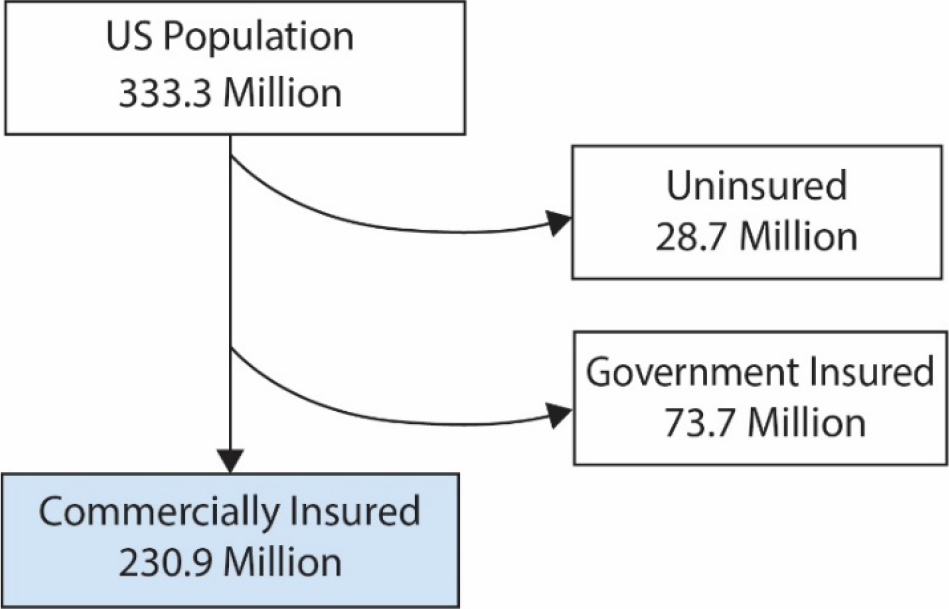
The number of people affected by commercial and Medicare Part D formularies in the U.S.

The 2022 market share for the two largest publicly available formularies, CVS Caremark Performance Standard Control (CVS) and Express Scripts National Preferred (ESI), is 33% and 24%, respectively.^13^

### Drug Classes Evaluated

We evaluated drug classes currently affected by CVS Health and ESI formulary exclusions utilized to treat cardiovascular, neurologic, and dermatologic conditions. We chose these drug classes since they all require therapeutic substitution due to the formulary exclusion policy. Therapeutic substitution occurs when an insurer or PBM mandates a patient to utilize a medicine that does not possess the drug’s active ingredient prescribed by their provider. Exclusion of treatments in all three drug classes forces discontinuation or substitution of therapy, leading to adverse patient outcomes.

## METHODS

### Data Sources

We used the CVS Health and ESI national formularies because both organizations publish a transparent national formulary and identify their excluded medicines.

We searched the published medical literature and the grey web to obtain data for our calculations and modeling of patient outcomes. We used the PubMed.gov and EMBASE databases to search published peer-reviewed medical literature. We searched the grey web with the Google search engine. For the prevalence of each indication evaluated, we used that indication(s) joined with the operator “AND” to “prevalence OR epidemiology” as search terms. For the proportion of people likely to discontinue treatment or have a related adverse event (medical consequence of discontinuation OR side effect of a new drug) after an exclusion, we used the indication(s) joined by “AND” to “nonmedical switching” as search terms. We limited all searches to the same 20-year period of 2002 to 2022 and completed all queries on March 31, 2023.

We obtained market share data of the three drug classes from IQVIA, which is a private company that owns proprietary databases that are utilized by many entities in the evaluation of the biopharmaceutical marketplace in the U.S. Within each class or subclass of medications evaluated, we calculated the market share for each medicine as the total number of prescriptions for that medication divided byall prescriptions for all drugs in the class or subclass evaluated.

### Calculating the Potential Effects of a Specific Exclusion

We calculated the number of people who could be affected by an exclusion as the number of individuals in the U.S. with commercial or Medicare Part D insurance (commercially insured lives) multiplied by the indication prevalence, then formulary market share, and then medication market share (Figure 2).^13^ We then calculated the number of people likely to discontinue therapy or have an adverse outcome due to an exclusion using our findings from the literature on discontinuation rates and adverse events (disease worsening from discontinuation plus adverse effects of switching to a new drug).

**Figure 2.**
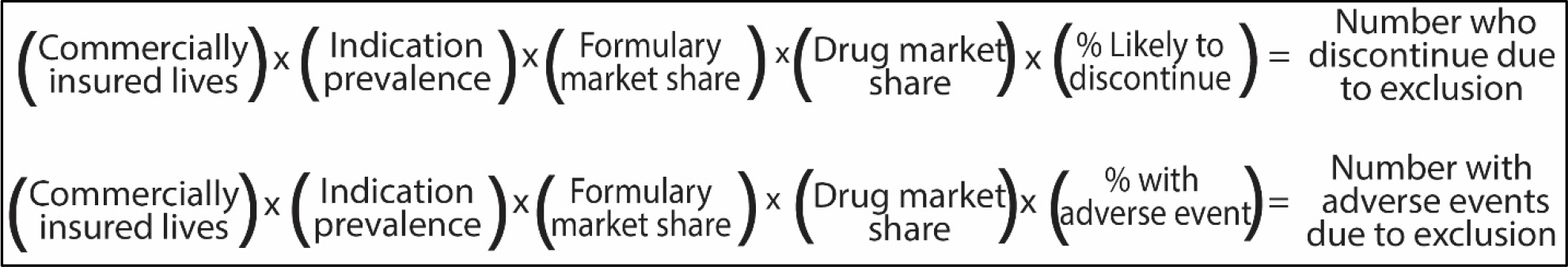
Calculation for the number of patients affected by specific exclusion in a specific formulary.

## RESULTS

### Disease Prevalence for Indications Evaluated

*Anticoagulants* are indicated for the treatment of atrial fibrillation (AFib) and/or venous thromboembolism (VTE), including pulmonary embolism (PE) and deep vein thrombosis (DVT). Epidemiologic studies show the age-adjusted prevalence of AFib in the U.S. is 1% to 2.9%.^14-16^ Nominal data is available on the prevalence of VTE, with a 2011 report of 0.4% to 0.5% in the U.S.^17^ Based on these data and the market share of the CVS and ESI formularies,^13^ 800,000 to 2.6 million individuals could be affected by the exclusion of an anticoagulant by CVS or ESI (Figure 3a).

**Figure 3.**
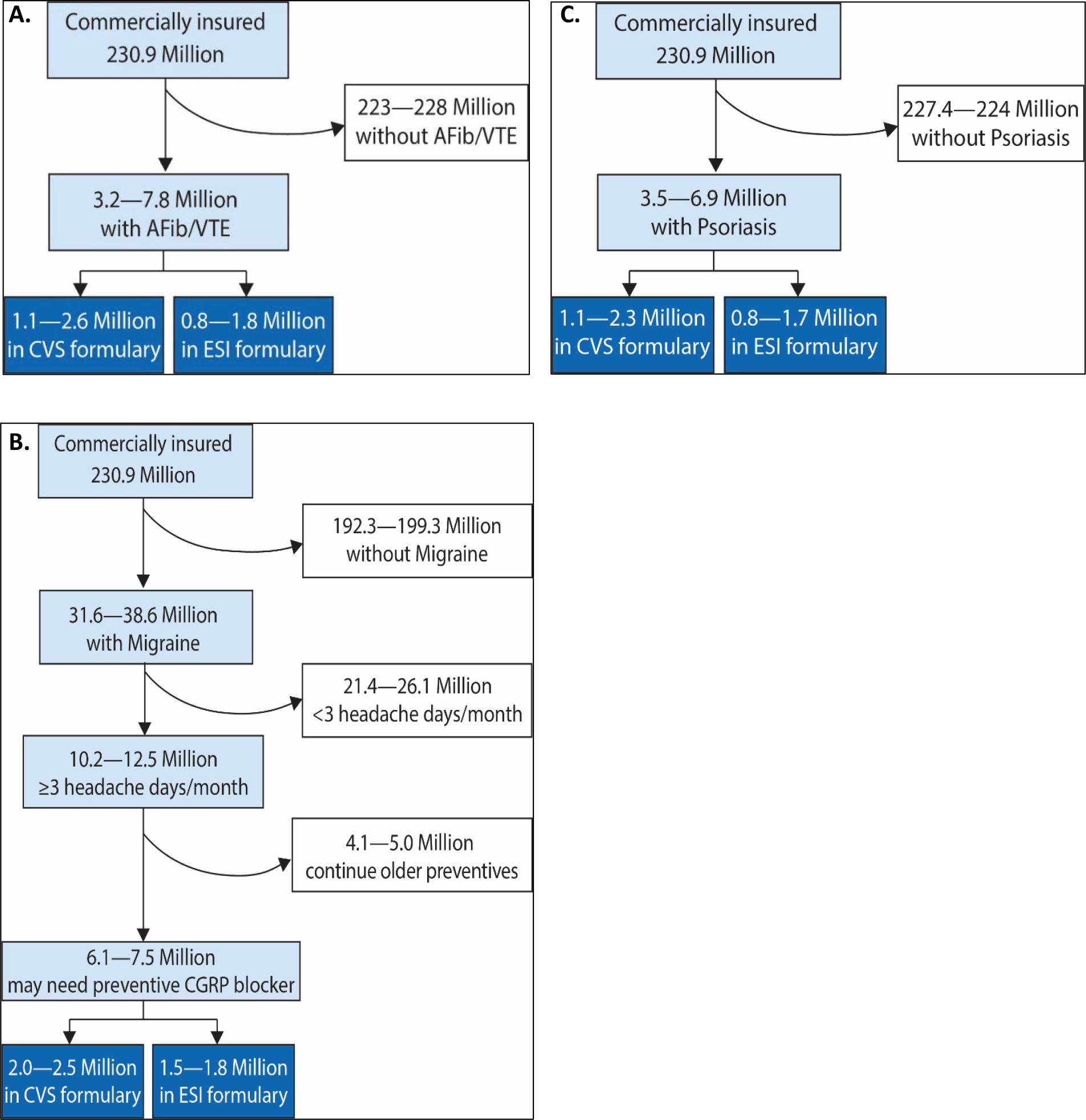
The number of people potentially affected by the exclusion of a brand-name anticoagulant (A), migraine preventive (B), or anti-psoriatic (C) medication.

*Migraine preventive treatments* are indicated for people with migraine who have more than three migraine attacks per month.^18^ The age-adjusted prevalence of migraine is 9.5% to 11.6% (∼31.6 to 38.6 million cases). Among people with migraine, 33.3% have more than three migraine attacks per month, making them eligible for preventive treatment.^19-21^ However, many eligible patients do not receive preventive treatment. Among those who do, only 40% continue the use of older medicines that do not block the activity of calcitonin gene-related peptide (CGRP) after six months.^22,23^ Based on these data and the market share of the CVS and ESI formularies, 1.5 million to 2.5 million individuals could be affected by excluding a migraine prevention treatment (Figure 3b) that blocks CGRP.

*Antipsoriatic agents* are indicated for people with topical psoriasis, for which prevalence in the U.S. has consistently been reported as 1.5% to 3% (∼3.5-6.9 million people).^24-26^ Based on these data and the market share of the CVS and ESI formularies,^13^ 800,000 to 2.3 million individuals could be affected by the exclusion of an antipsoriatic medication by CVS or ESI (Figure 3c).

### Rates of Discontinuation and Adverse Events After Switching

#### Anticoagulants

Studies show that 17% to 30% of patients discontinue all anticoagulant treatment upon exclusion of apixaban from a national formulary.^27,28^ Using this discontinuation rate, up to 389,000 and 535,000 patients, respectively, would likely discontinue treatment with any anticoagulant if excluded by ESI or CVS. For those who discontinue anticoagulation therapy, the likelihood of stroke or other cardiovascular events increases by 45% to 85.^29^ In addition, 2% - 10% of patients who switched from anticoagulant medicines had an adverse event.^30-33^ We added the number of people likely to have a serious cardiovascular event (45% to 85% of the 17% to 30% who discontinue) to the number of people likely to have an adverse event after switching anticoagulants (2% to 10% of the 83% who switch). This calculation shows that up to 422,000 and 580,000 patients may have an adverse event upon forced switching due to CVS Health and ESI formulary exclusions, respectively (Table 1a and 1b).

**Table 1a.** Potential Effects of CVS Excluding a Brand-Name Anticoagulant (000s)

|  |  |  |  |  |
| --- | --- | --- | --- | --- |
| Drug | Market share | Potentially affected | Likely to discontinue | Adverse events |
| Eliquis (apixaban) | 68.88% | 734-1,784 | 125-535 | 68-580 |
| Xarelto (rivaroxaban) | 29.76% | 317-771 | 54-231 | 30-251 |
| Pradaxa (dabigatran) | 1.31% | 14-34 | 2.3-10 | 1.3-11 |
| Savaysa (edoxaban) | 0.06% | 0.6-1.5 | 0.1-0.5 | 0.06-0.5 |

**Table 1b.** Potential Effects of ESI Excluding a Brand-Name Anticoagulant (000s)

|  |  |  |  |  |
| --- | --- | --- | --- | --- |
| Drug | Market share | Potentially affected | Likely to discontinue | Adverse events |
| Eliquis (apixaban) | 68.88% | 534-1,298 | 91-389 | 50-422 |
| Xarelto (rivaroxaban) | 29.76% | 231-561 | 39-168 | 21-182 |
| Pradaxa (dabigatran) | 1.31% | 10-25 | 1.7-7.4 | 0.9-8.0 |
| Savaysa (edoxaban) | 0.06% | 0.4-1.1 | 0.08-0.3 | 0.04-.4 |

#### Migraine preventive agents

Data regarding the implications of discontinuing or switching CGRP medicines due to formulary exclusions are limited. However, a meta-analysis of forced non-equivalent substitutions across multiple drug classes showed 9% to 19% of patients discontinue treatment.^34^ Based on these data, we calculated that a maximum of 374,000 individuals are likely to discontinue therapy due to a formulary exclusion by CVS Health or ESI. Regarding adverse events related to discontinuation, one published study documented increased migraine attack frequency after stopping a monoclonal antibody CGRP blocker for any reason.^35^ Another study showed stopping or switching any migraine preventive treatment increased migraine attack frequency and/or serious adverse events in at least half of patients.^36^ Based on these data, we calculated that 50% of patients, up to 986,000 people, would have worsening disease and other adverse events after the exclusion of a migraine preventive agent by ESI or CVS (Table 2a and 2b).

**Table 2a.**
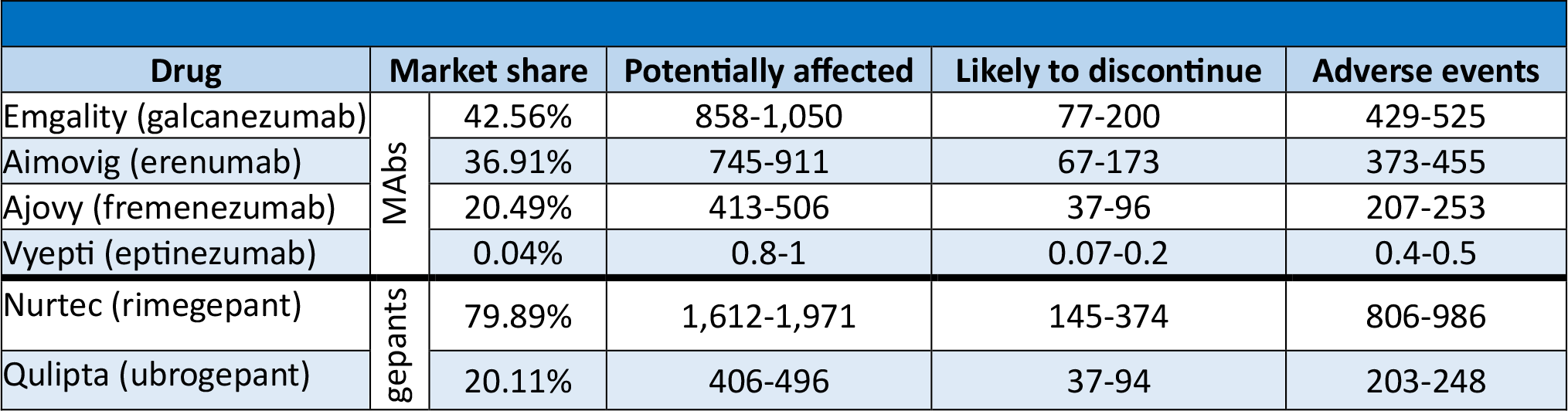
Potential Effects of CVS Excluding a Brand-Name Migraine Preventive Treatment (000s)

**Table 2b.** Potential Effects of ESI Excluding a Brand-Name Migraine Preventive Treatment (000s)

|  |  |  |  |  |  |
| --- | --- | --- | --- | --- | --- |
| Drug |  | Market share | Potentially affected | Likely to discontinue | Adverse events |
| Emgality (galcanezumab) | MAbs | 42.56% | 625-764 | 56-145 | 312-382 |
| Aimovig (erenumab) |  | 36.91% | 542-662 | 49-103 | 271-331 |
| Ajovy (fremenezumab) |  | 20.49% | 301-368 | 27-70 | 150-184 |
| Vyepti (eptinezumab) |  | 0.04% | 0.6-0.7 | 0.05-0.14 | 0.3-0.4 |
| Nurtec (rimegepant) | gepants | 79.89% | 1,172-1,433 | 106-272 | 586-717 |
| Qulipta (ubrogepant) |  | 20.11% | 295-361 | 27-69 | 148-180 |

#### Anti-psoriatic agents

A meta-analysis showed that 6% - 9% of patients discontinue therapy upon being forced to switch tumor necrosis factor inhibitors (TNFi).^37^ For other anti-psoriatic medications, we used the 9% to 19% discontinuation rate seen in a meta-analysis of non-medical switching across multiple drug classes^34^ and in studies of non-medical switching of biological medications.^38-40^ The rate of adverse events from all of these agents is high, and studies show that switching results in 15% to 35% of patients having adverse events.^41^ Using these data, we calculate up to 28,000 patients may discontinue therapy, and up to 311,000 may have an adverse event because of a formulary exclusion (Tables 3a and 3b).

**Table 3a.** Potential Effects of CVS Excluding a Brand-Name or Biosimilar Psoriasis Treatment (000s)

|  |  |  |  |  |  |
| --- | --- | --- | --- | --- | --- |
| Drug |  | Market share | Potentially affected | Likely to discontinue | Adverse events |
| Humira (adalimumab) | TNF inhibitors | 38.92% | 445-890 | 27-80 | 67-311 |
| Enbrel (etanercept) |  | 13.18% | 151-301 | 9-27 | 23-105 |
| Cimzia (certolizumab) |  | 2.50% | 29-57 | 2-5 | 4-20 |
| Simponi (golimumab) |  | 1.41% | 16-32 | 1-3 | 2-11 |
| Remicade (infliximab) |  | 0.52% | 6-12 | 0.4-1 | 0.9-4 |
| Inflectra (infliximab) |  | 0.30% | 3-7 | 0.2-0.6 | 0.2-2 |
| Renflexis (infliximab) |  | 0.03% | 0.3-0.7 | 0.02-0.06 | 0.05-0.2 |
| Avsola (infliximab) |  | 0.02% | 0.2-0.5 | 0.01-0.04 | 0.03-0.1 |
| Infliximab (infliximab) |  | 0.02% | 0.2-0.5 | 0.01-0.04 | 0.03-0.1 |
| Otezla (apremilast) | Other | 8.55% | 98-195 | 9-19 | 15-68 |
| Cosentyx (ustekinumab) |  | 7.30% | 83-167 | 8-32 | 13-58 |
| Stelara (ustekinumab) |  | 6.38% | 73-146 | 7-28 | 11-51 |
| Taltz (ixekizumab) |  | 6.17% | 71-141 | 6-27 | 11-49 |
| Xeljanz (tofacitinib) |  | 5.24% | 60-120 | 5-23 | 9-42 |
| Tremfya (guselkumab) |  | 3.46% | 40-79 | 4-15 | 6-28 |
| Rinvoq(upadacitinib) |  | 3.23% | 37-74 | 3-14 | 6-26 |
| Skyrizi (risankizumab) |  | 2.63% | 30-60 | 3-11 | 5-21 |
| Siliq (brodalumab) |  | 0.07% | 0.8-1.6 | 0.07-0.3 | 0.1-0.6 |
| Ilumya (tildrakizumab) |  | 0.06% | 0.7-1.4 | 0.06-0.3 | 0.1-0.5 |

**Table 3b.**
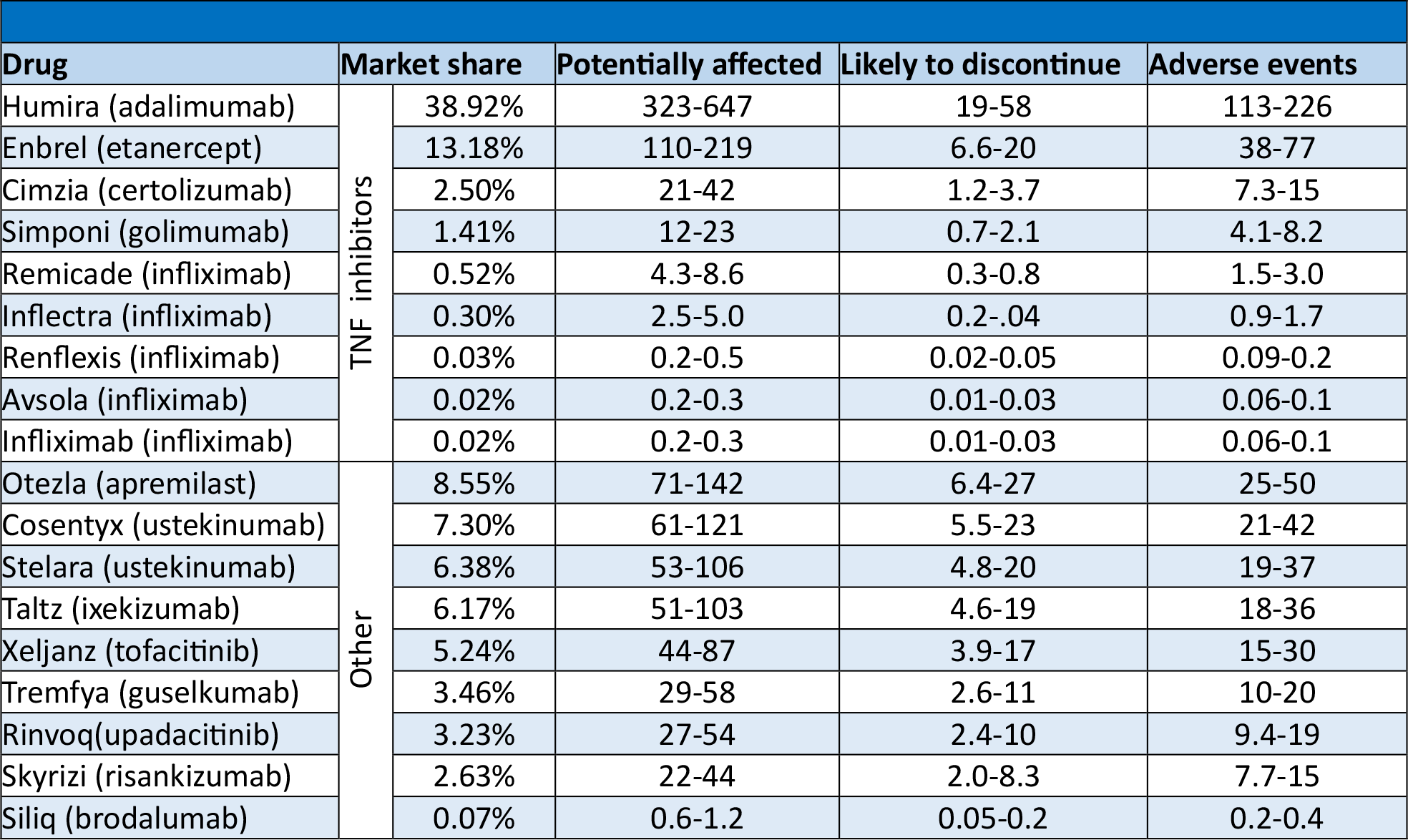
Potential Effects of ESI Excluding a Brand-Name or Biosimilar Psoriasis Treatment (000s)

## DISCUSSION

The purpose of formularies has evolved from excluding more expensive brand-name drugs in favor of equivalent generic medicines to maximizing rebates and discounts collected as profit by PBMs. Such business tactics ultimately lead to questionable decision-making when developing formularies that do not benefit patients. In fact, over half of the exclusions in the second-largest PBM in the U.S. may cause economic or medical harm to the patient.^5,9^

For this study, we examined three therapeutic areas with current exclusions of medicines. Our model is based on the number of people in the U.S. receiving benefits through CVS Health and ESI who may be utilizing one of the medicines identified in each drug class. Applying published discontinuation rates and adverse events after forced switching, we have calculated the number of patients who could be affected by excluding these agents.

Although the number of people affected is small for some medicines with small market shares, the total number of people who could be harmed by excluding anticoagulants, migraine preventatives, or antipsoriatic drugs is substantial. Excluding the most commonly used anticoagulant, for example, could increase the number of strokes by 227,000 to 459,000 people in six months. Exclusion of migraine preventive treatments by both formularies would likely worsen the frequency of migraine attacks, thus increasing disability for as many as 875,000 patients. Finally, excluding anti-psoriatic drugs could cause serious side effects for over 500,000 patients.

It is also important to note that the calculated increases in untreated disease and adverse events carry costs often not factored into equations of medication cost-effectiveness. In addition, whether any of the substituted drugs are truly more or less cost-effective cannot be determined in the current system in which actual drug net prices of the medicines are not transparent. What is certain, however, is that there are costs of forcing patients to switch to a non-equivalent drug that arises from untreated disease and increased adverse events. Those costs warrant further investigation.

### LIMITATIONS

Data on how many people use a particular drug for a specific indication are limited, and the annual effects of medication switching can occur over many years. In some cases, we also had to utilize non-specific and generalized prevalence data concerning discontinuation and switching. We modeled what would happen in the two largest formularies, but hundreds of unique formularies also exist, and each may have specific exclusions based on individual contracts negotiated. However, the data used to model the number of patients who could be affected are based on the most extensive available data sets. Thus, they can be considered an appropriate proxy for all formularies.

## CONCLUSIONS

Evaluating the actual costs and benefits of formulary exclusions in the current U.S. drug pricing environment is impossible when net prices paid by insurers and PBMs are not public. The unknown specific costs of forced switching further hamper the ability of economists and policymakers to perform cost-benefit analyses. We know that excluding medicines impacts hundreds of thousands of patients and increases morbidity and mortality. Forced drug substitution due to formulary exclusions increases the burden of illness on patients and their care partners. It also increases costs for the ultimate payers in the healthcare system, patients, caregivers, employers, and the government. Determining these costs should be part of cost-benefit analyses of any formulary exclusion.

## Data Availability

All data produced in the present study are available upon reasonable request to the authors

## REFERENCES

1. Kelly D. A brief history of drug formularies and what to expect in 2019. CoverMyMeds. 2018. Accessed October 3, 2023. https://www.covermymeds.com/main/insights/articles/a-brief-history-of-drug-formularies-and-what-to-expect-in-2019/

2. Gilman BH, Kautter J. Impact of multitiered copayments on the use and cost of prescription drugs among Medicare beneficiaries. Health Serv Res. 2008;43:478–495.

3. U.S. Department of Health and Human Services. Are generic drugs the same as brand name drugs? 2014. Accessed October 3, 2023. https://www.hhs.gov/answers/public-health-and-safety/are-generic-drugs-the-same-as-brand-name-drugs/index.html

4. Alston M, Dieguez G, Tomicki S. A primer on prescription drug rebates: Insights into why rebates are a target for reducing prices. Milliman.2018. Accessed October 3, 2023. https://www.milliman.com/en/insight/a-primer-on-prescription-drug-rebates-insights-into-why-rebates-are-a-target-for-reducing

5. Popovian R, Sydor A, Pitts P. Analysis of drug formulary exclusions from the patient’s perspective. Health Science Journal. 2022;16(3):1–4. doi:10.36648/1791-809X.16.4.937

6. Sood N, Ribero R, Ryan M, Van Nuys K. The association between drug rebates and list prices. University of Southern California, Leonard D. Schaeffer Center for Health Policy and Economics. 2020. Accessed October 3, 2023. https://healthpolicy.usc.edu/research/the-association-between-drug-rebates-and-list-prices/

7. Lakdawalla D, Li M. Association of drug rebates and competition with out-of-pocket co-insurance in Medicare Part D, 2014 to 2018. JAMA Netw Open. 2021;4(5):e219030. doi:10.1001/jamanetworkopen.2021.9030

8. Texas Department of Health Insurance. Life, health and accident insurance reports, prescription drug cost transparency, pharmacy benefit managers. Accessed October 3, 2023. https://www.tdi.texas.gov/reports/report3.html

9. Crea, Sydor, and Popovian. 2023 Update: Analysis of drug formulary exclusions from the patient’s perspective. Manuscript in preparation. 2023.

10. United States Census Bureau. Quick Facts Table. Accessed October 3, 2023. https://www.census.gov/quickfacts/fact/table

11. Kaiser Family Foundation. Health Insurance Coverage of the Total Population. 2021. Accessed July 5, 203. https://www.kff.org/other/state-indicator/total-population/

12. Kaiser Family Foundation. An Overview of the Medicare Part D Prescription Drug Benefit. October 19, 2022. Accessed July 5, 2023. https://www.kff.org/medicare/fact-sheet/an-overview-of-the-medicare-part-d-prescription-drug-benefit/

13. Fein A. The big three PBMs’ 2023 formulary exclusions: observations on insulin, Humira, and biosimilars. Drug Channels. January 10, 2023. Accessed July 12, 2023 https://www.drugchannels.net/2023/01/the-big-three-pbms-2023-formulary.html

14. Colilla S, Crow A, Petkun W, Singer DE, Simon T, Liu X. Estimates of current and future incidence and prevalence of atrial fibrillation in the U.S. adult population. Am J Cardiol. 2013;112(8):1142–1147. doi:10.1016/j.amjcard.2013.05.063

15. Go AS, Hylek EM, Phillips KA, et al. Prevalence of diagnosed atrial fibrillation in adults: national implications for rhythm management and stroke prevention: the Anticoagulation and Risk Factors in Atrial Fibrillation (ATRIA) Study. JAMA. 2001;285(18):2370–2375. doi:10.1001/jama.285.18.2370

16. Benjamin EJ, Muntner P, Alonso A, et al. Heart Disease and Stroke Statistics-2019 Update: A Report From the American Heart Association [published correction appears in Circulation. 2020 Jan 14;141(2):e33]. Circulation. 2019;139(10):e56–e528. doi:10.1161/CIR.0000000000000659

17. Deitelzweig SB, Johnson BH, Lin J, Schulman KL. Prevalence of clinical venous thromboembolism in the USA: current trends and future projections. Am J Hematol. 2011;86(2):217–220. doi:10.1002/ajh.21917

18. Ailani J, Burch RC, Robbins MS; Board of Directors of the American Headache Society. The American Headache Society Consensus Statement: Update on integrating new migraine treatments into clinical practice. Headache. 2021;61(7):1021–1039. doi:10.1111/head.14153

19. Burch R, Rizzoli P, Loder E. The prevalence and impact of migraine and severe headache in the United States: Updated age, sex, and socioeconomic-specific estimates from government health surveys. Headache. 2021;61(1):60–68. doi:10.1111/head.14024

20. GBD 2016 Headache Collaborators. Global, regional, and national burden of migraine and tension-type headache, 1990-2016: a systematic analysis for the Global Burden of Disease Study 2016 [published correction appears in Lancet Neurol. 2021 Dec;20(12):e7]. Lancet Neurol. 2018;17(11):954–976. doi:10.1016/S1474-4422(18)30322-3

21. Buse DC, Reed ML, Fanning KM, Bostic RC, Lipton RB. Demographics, headache features, and comorbidity profiles in relation to headache frequency in people with migraine: results of the American Migraine Prevalence and Prevention (AMPP) Study. Headache. 2020;10.1111/head.13966. doi:10.1111/head.13966

22. Blumenfeld AM, Bloudek LM, Becker WJ, et al. Patterns of use and reasons for discontinuation of prophylactic medications for episodic migraine and chronic migraine: results from the second international burden of migraine study (IBMS-II). Headache. 2013; 53(4): 644–655. doi:10.1111/head.12055

23. Bonafede M, McMorrow D, Noxon V, Desai P, Sapra S, Silberstein S. Care among migraine patients in a commercially insured population. Neurol Ther. 2020;9(1):93–103. doi:10.1007/s40120-020-00179-3

24. Parisi R, Iskandar IYK, Kontopantelis E, et al. National, regional, and worldwide epidemiology of psoriasis: systematic analysis and modeling study. BMJ. 2020;369:m1590. doi:10.1136/bmj.m1590

25. Armstrong AW, Mehta MD, Schupp CW, Gondo GC, Bell SJ, Griffiths CEM. Psoriasis prevalence in adults in the United States. JAMA Dermatol. 2021;157(8):940–946. doi:10.1001/jamadermatol.2021.2007

26. Kurd SK, Gelfand JM. The prevalence of previously diagnosed and undiagnosed psoriasis in U.S. adults: results from NHANES 2003-2004 [published correction appears in J Am Acad Dermatol. 2009;61(3):507]. J Am Acad Dermatol. 2009;60(2):218–224. doi:10.1016/j.jaad.2008.09.022

27. Deitelzweig S, Terasawa E, Atreja N, et al. Payer formulary tier increases of apixaban: how patients respond and potential implications. Curr Med Res Opin. 2023;39(8):1093–1101. doi:10.1080/03007995.2023.2232636

28. Deitelzweig S, Terasawa E, Kang A, et al. Payer formulary exclusions of apixaban: how patients respond and potential implications. Curr Med Res Opin. 2022;38(11):1885–1890. doi:10.1080/03007995.2022.2128189

29. Rivera-Caravaca JM, Roldán V, Esteve-Pastor MA, et al. Cessation of oral anticoagulation is an important risk factor for stroke and mortality in atrial fibrillation patients. Thromb Haemost. 2017;117(7):1448–1454. doi:10.1160/TH16-12-0961

30. Eliquis. Prescribing information. Bristol Myers Squibb. New York, NY. Accessed September 29, 2023. https://packageinserts.bms.com/pi/pi_eliquis.pdf

31. Xarelto. Prescribing information. Janssen. Titusville, NJ. Accessed September 29, 2023. https://www.janssenlabels.com/package-insert/product-monograph/prescribing-information/XARELTO-pi.pdf

32. Pradaxa. Prescribing information. Boehringer Ingelheim. Ridgefield, CT. Accessed September 29, 2023. https://www.accessdata.fda.gov/drugsatida_docs/label/2011/022512s007lbl.pdf

33. Savaysa. Prescribing information. Daiichi Sankyo, Inc. Parsippany, NJ. Accessed October 3, 2023. https://www.accessdata.fda.gov/drugsatida_docs/label/2015/206316lbl.pdf

34. Weeda ER, Nguyen E, Martin S, et al. The impact of non-medical switching among ambulatory patients: an updated systematic literature review. J Mark Access Health Policy. 2019;7(1):1678563. doi:10.1080/20016689.2019.1678563

35. Vernieri F, Brunelli N, Messina R, et al. Discontinuing monoclonal antibodies targeting CGRP pathway after one-year treatment: an observational longitudinal cohort study. J Headache Pain. 2021;22(1):154. doi:10.1186/s10194-021-01363-y

36. Hepp Z, Dodick DW, Varon SF, et al. Persistence and switching patterns of oral migraine prophylactic medications among patients with chronic migraine: a retrospective claims analysis. Cephalalgia. 2017;37(5):470–485. doi: 10.1177/0333102416678382

37. Liu Y, Skup M, Yang M, Qi CZ, Wu EQ. Discontinuation and switchback after non-medical switching from originator tumor necrosis factor alpha (TNF) inhibitors to biosimilars: a meta-analysis of realworld studies from 2012 to 2018 [published correction appears in Adv Ther. 2022 Nov;39(11):5300]. Adv Ther. 2022;39(8):3711–3734. doi:10.1007/s12325-022-02173-7

38. Aceves-Ávila FJ, Hernández Vásquez JR, Sicsick S, et al. Not the same, but is it the same? Cycling of biologic agents in rheumatoid arthritis. Experience in the Instituto Mexicano del Seguro Social. Reumatol Clin (Engl Ed). 2022;18(6):361–367. doi:10.1016/j.reumae.2021.02.004

39. Chambers JD, Rane PB, Neumann PJ. The impact of formulary drug exclusion policies on patients and healthcare costs. Am J Manag Care. 2016;22(8):524–531. https://www.ajmc.com/view/the-impactof-formulary-drug-exclusion-policies-on-patients-and-healthcare-costs

40. Melville AR, Md Yusof MY, Fitton J, et al. Real-world experience of effectiveness of non-medical switch from originator to biosimilar rituximab in rheumatoid arthritis. Rheumatology (Oxford). 2021;60(8):3679–3688. doi:10.1093/rheumatology/keaa834

41. Tweehuysen L, van den Bemt BJF, van Ingen IL, et al. Subjective complaints as the main reason for biosimilar discontinuation after open-label transition from reference infliximab to biosimilar infliximab. Arthritis Rheumatol. 2018;70(1):60–68. doi:10.1002/art.40324

